## Supplementary info for "Artificial intelligence-based histopathology image analysis identifies a novel subset of endometrial cancers with distinct genomic features and unfavourable outcome"

### Supplementary Materials

#### List of Figures

**Supplementary Fig. 1:** ROC plots of the trained VarMIL networks in the (A) discovery and (B) validation sets.
**Supplementary Fig. 2:** DSS and PFS KM curves associated with NSMP, *p53abn-like NSMP*, and p53abn cases in the (A) discovery set and the (B) validation set. The reported p-values compare the significance between *p53abn-like NSMP* and p53abn.
**Supplementary Fig. 3:** DSS and PFS KM curves associated with *NSMP-like p53abn* and p53abn cases in the (A) discovery set and the (B) validation set. *NSMP-like p53abn* represents cases that are p53abn as assessed by IHC but classified as NSMP based on H&E slides by the AI model.
**Supplementary Figure 4:** Performance benchmarking of Vanilla (A), IDaRS (B), Histogram (C), DeepMIL (D), and VLAD (E) models for the discovery and validation sets.
**Supplementary Figure 5:** Kaplan Meier curves along associated with the p53abn-like NSMP and NSMP groups for the discovery (A) and validation (B) cohorts using various deep learning frameworks. Note, DSS was not available for the TCGA part of the discovery cohort.
**Supplementary Fig. 6:** An overview of AI tumor-normal classifier and automatic annotation. (A) Extracting tumor and stroma patches from manually annotated slide and training a binary deep model. Red regions show annotated tumor regions, while green depicts stroma sections. (B) Extracting all patches from un-annotated slides and feeding them to the trained model to highlight only the tumor ones. The final tumor regions are shown by red contours.

#### List of Tables

**Supplementary Table 1:** Overview of cohorts.
**Supplementary Table 2:** Accuracy and other performance measures of the binary tumor-stroma classifier.
**Supplementary Table 3:** Performance metrics of the deep learning model for p53abn vs. NSMP classifier. The results are based on mean ± std of 10 cross-validation splits.
**Supplementary Table 4:** Detailed accuracy and other performance measures of the VarMIL network.
**Supplementary Table 5:** Statistics of NSMP and *p53abn-like NSMP* patients based on p53abn vs. NMSP classifiers.
**Supplementary Table 6:** Performance benchmarking of Vanilla (A), IDaRS (B), Histogram (C), DeepMIL (D), and VLAD (E) models for the discovery and validation sets.
**Supplementary Table 7:** Statistics of NSMP and p53abn-like NSMP patients based on p53abn vs. NMSP Vanilla (A), IDaRS (B), Histogram (C), DeepMIL (D), and VLAD (E) models for the discovery and validation sets.
**Supplementary Table 8:** Clinicopathologic features of the *p53abn-like NSMP* group in the discovery set.
**Supplementary Table 9:** Clinicopathologic features of the *p53abn-like NSMP* group in the validation set without excluding any patients.
**Supplementary Table 10:** Multi-variate Cox regression analysis showing the prognostic significance of *p53abn-like NSMP* group for PFS.

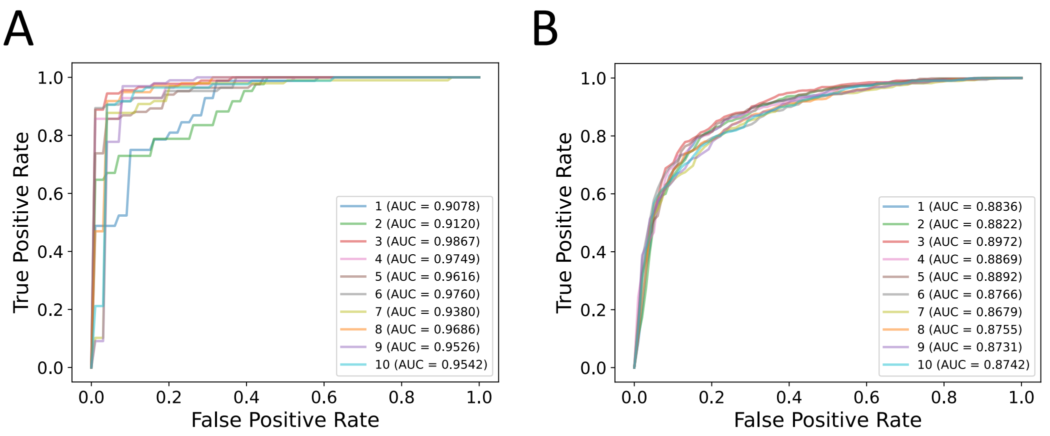

**Supplementary Fig. 1: ROC plots of the trained VarMIL networks in the (A) discovery and (B) validation sets.**

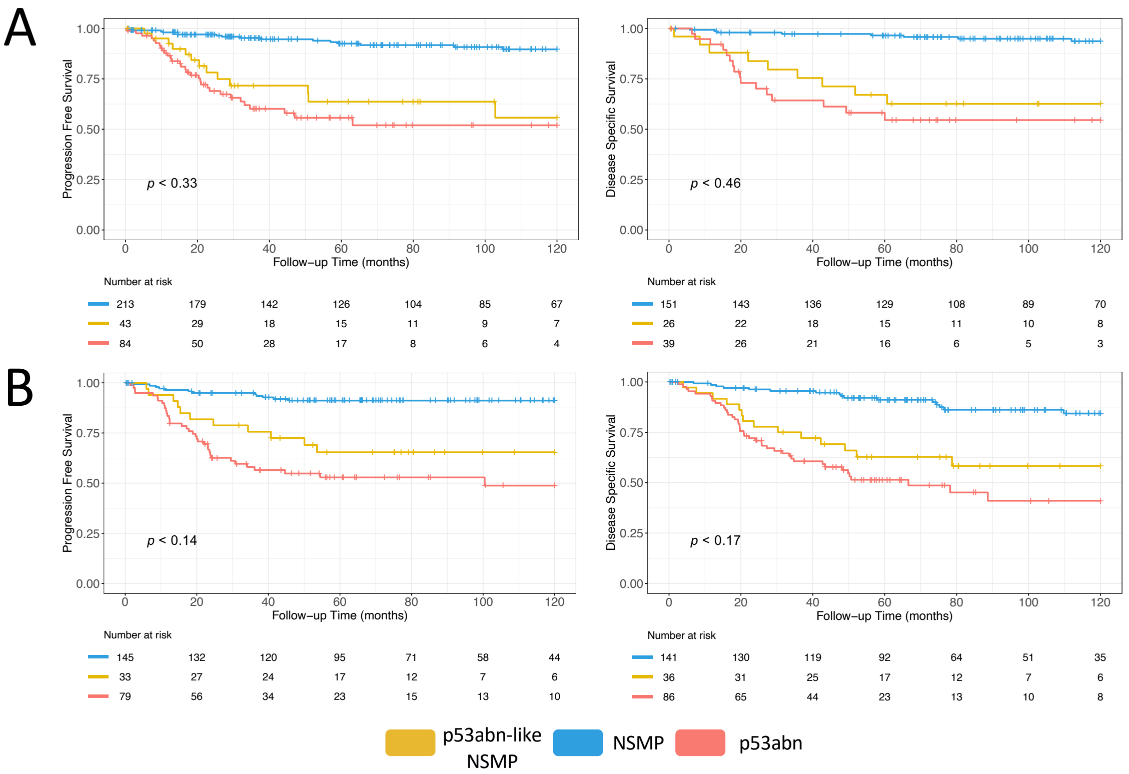

**Supplementary Fig. 2**: DSS and PFS KM curves associated with NSMP, *p53abn-like NSMP*, and p53abn cases in the (A) discovery set and the (B) validation set. The reported p-values compare the significance between *p53abn-like NSMP* and p53abn.

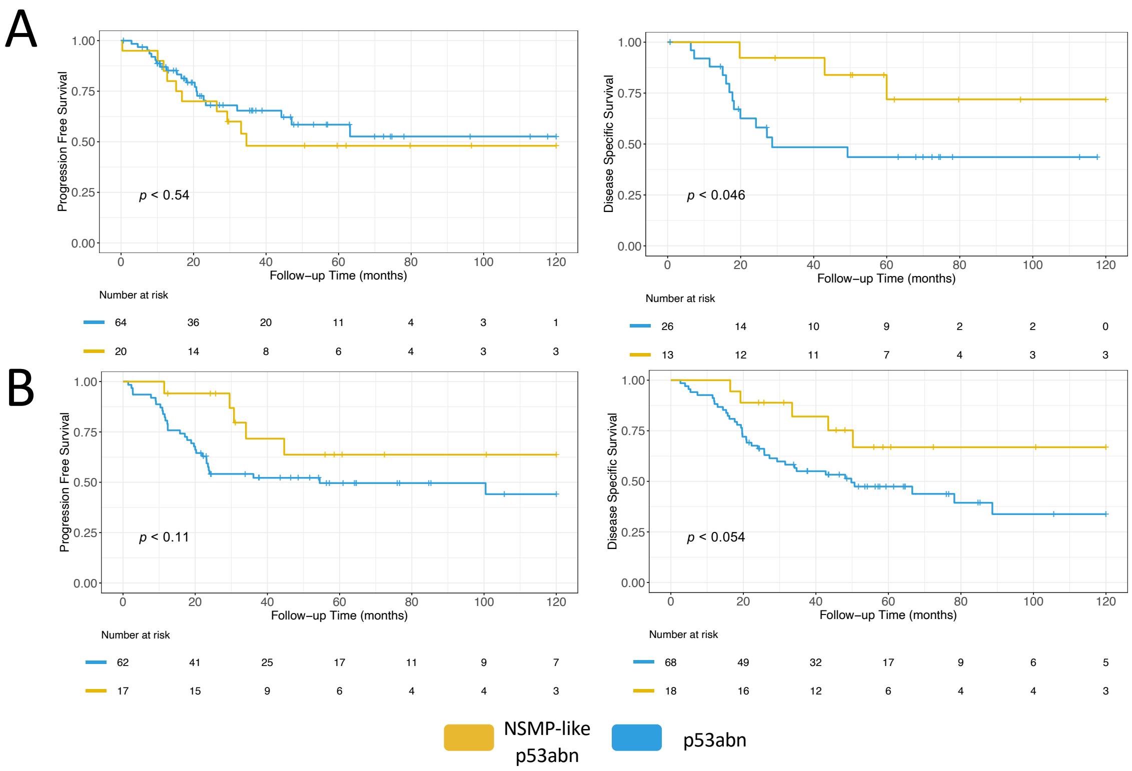

**Supplementary Fig. 3:** DSS and PFS KM curves associated with *NSMP-like p53abn* and p53abn cases in the (A) discovery set and the (B) validation set. *NSMP-like p53abn* represents cases that are p53abn as assessed by IHC but classified as NSMP based on H&E slides by the AI model.

A: Vanilla

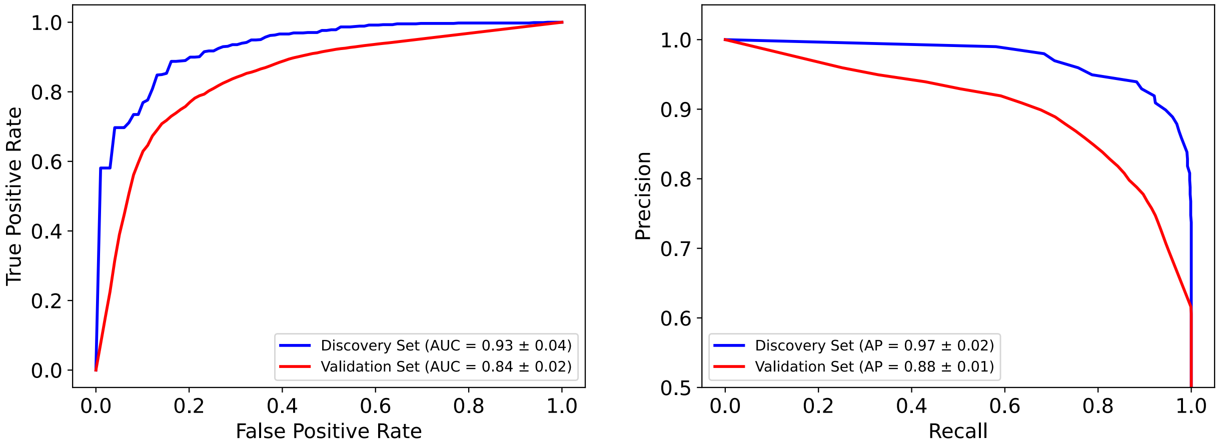

B: IDaRS

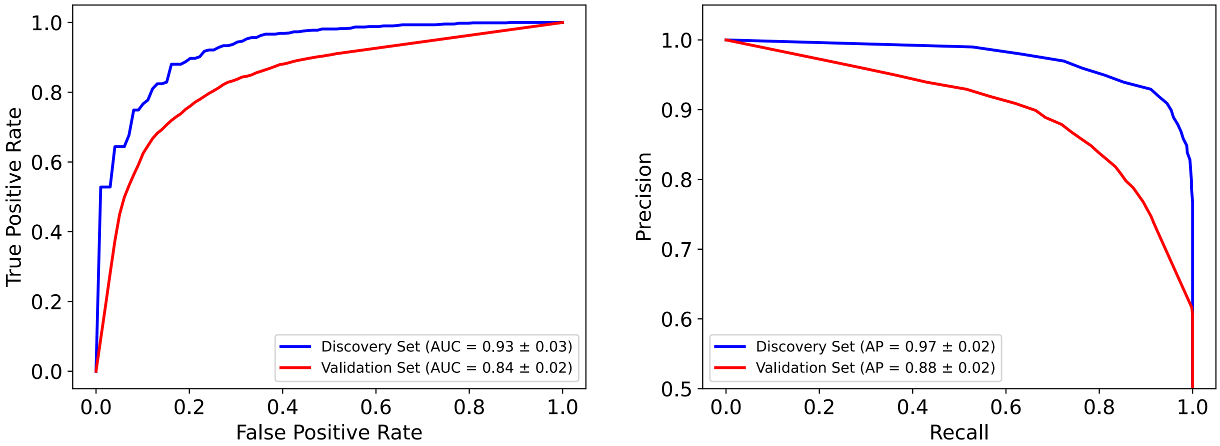

C: Histogram

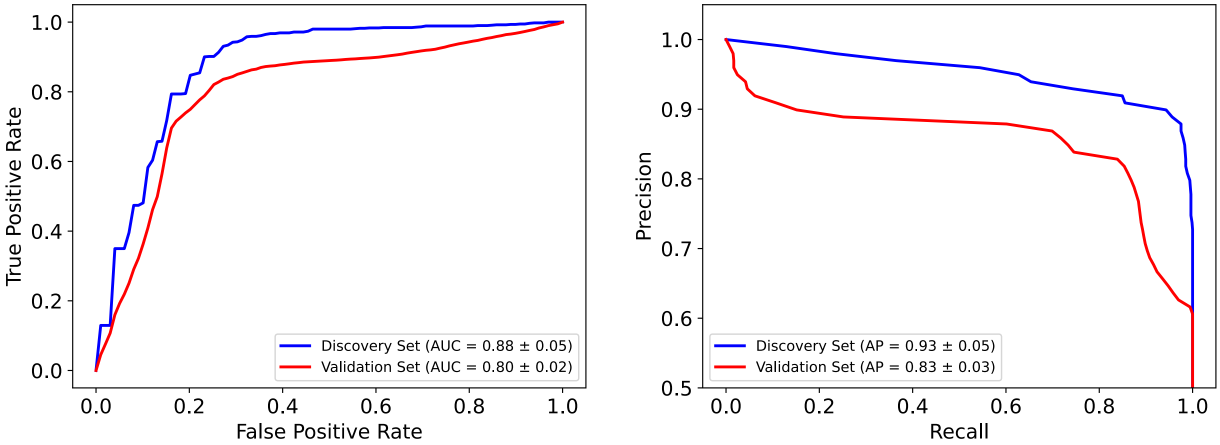

D: DeepMIL

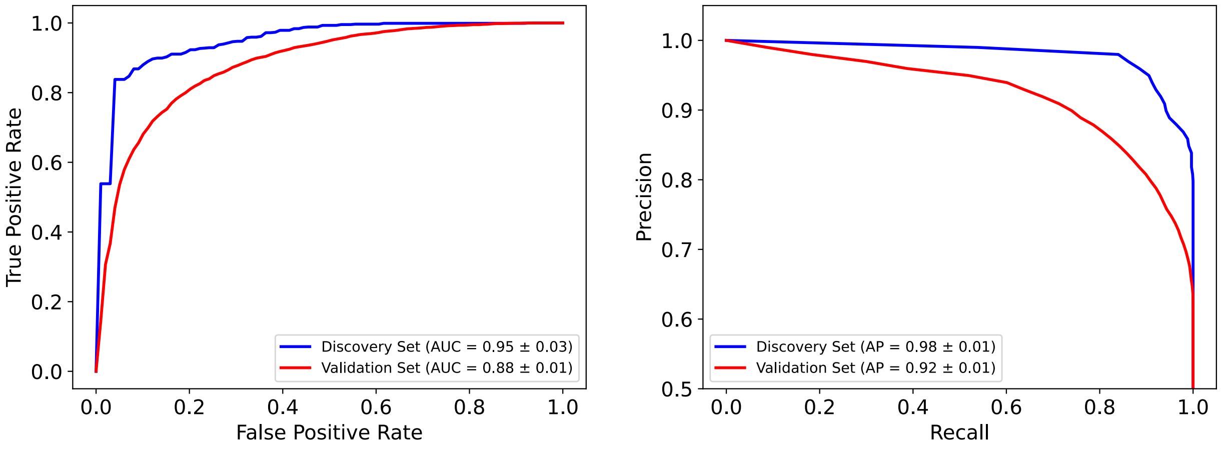

E: VLAD

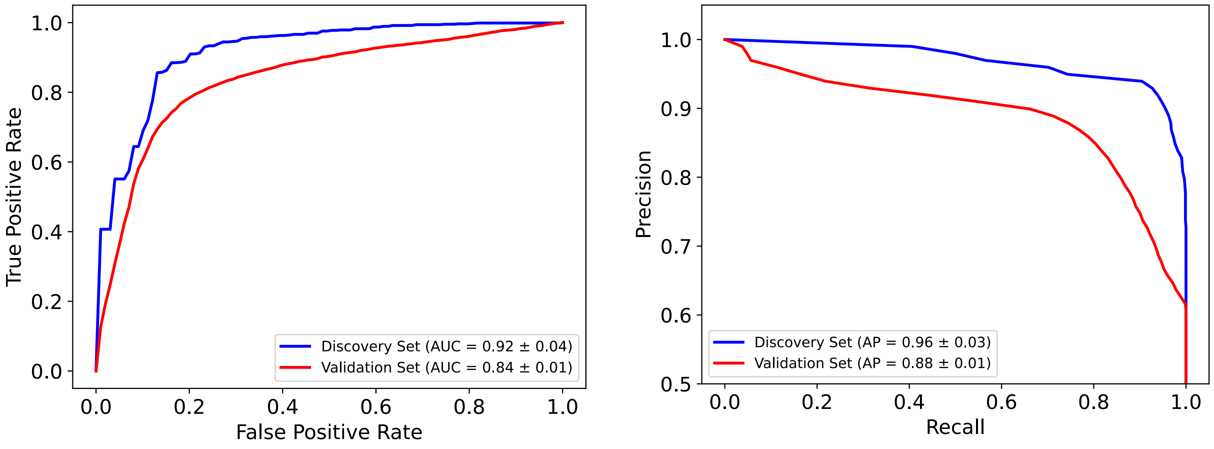

**Supplementary Figure 4:** Performance benchmarking of Vanilla (A), IDaRS (B), Histogram (C), DeepMIL (D), and VLAD (E) models for the discovery and validation sets.

**A: Discovery cohort**

**DSS** **PFS**

**Vanilla**

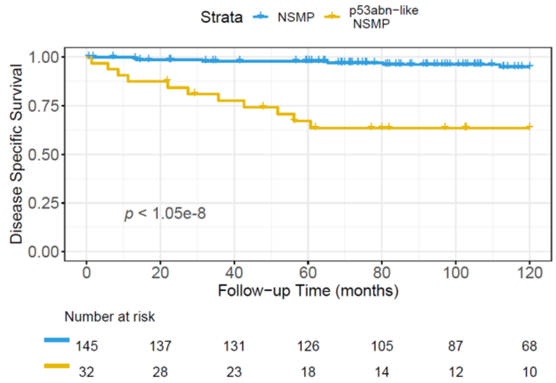

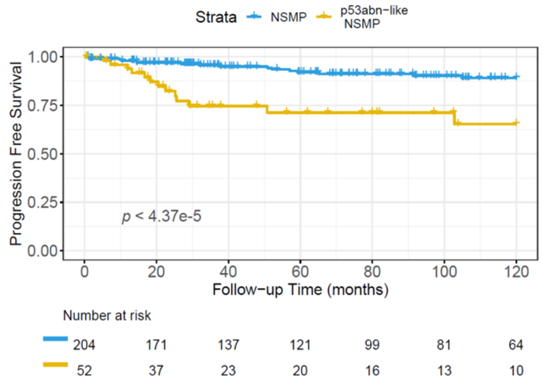

**IDaRS**

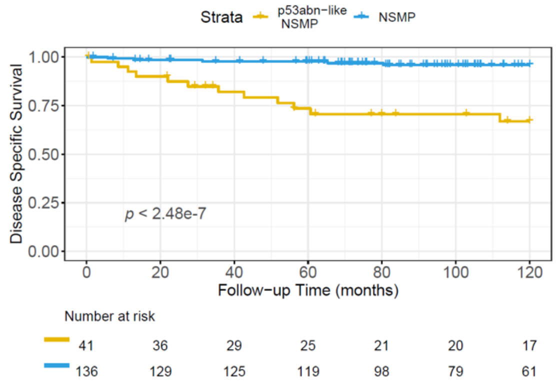

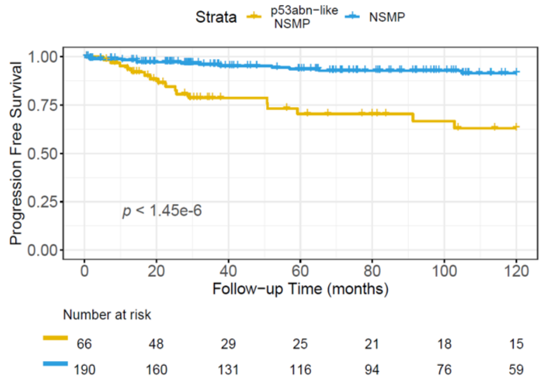

**Histogram-based**

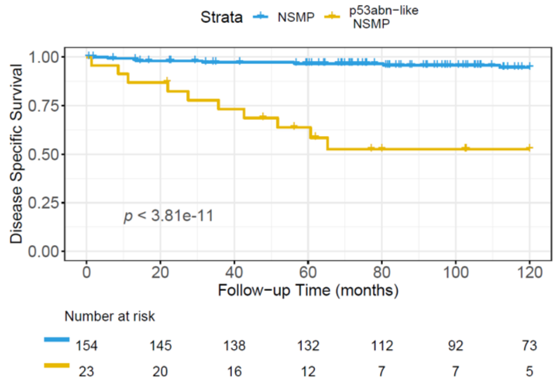

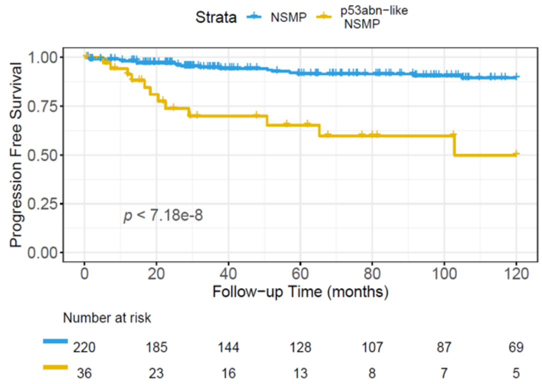

**DeepMIL**

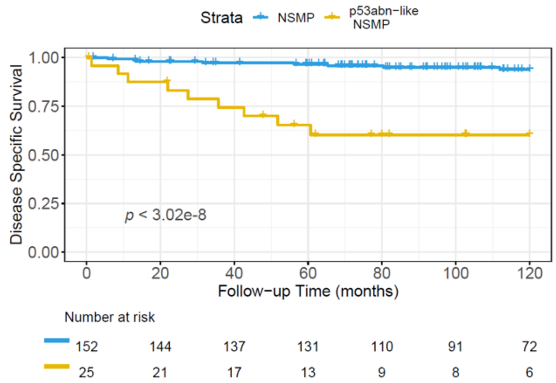

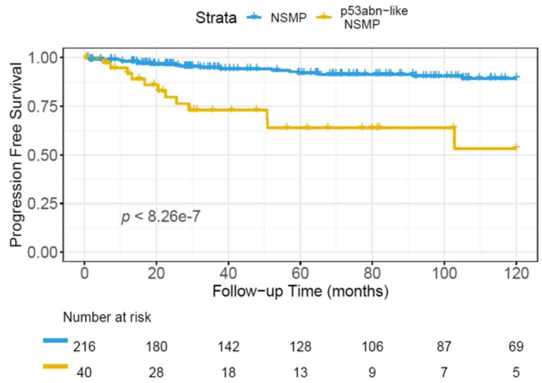

**VLAD**

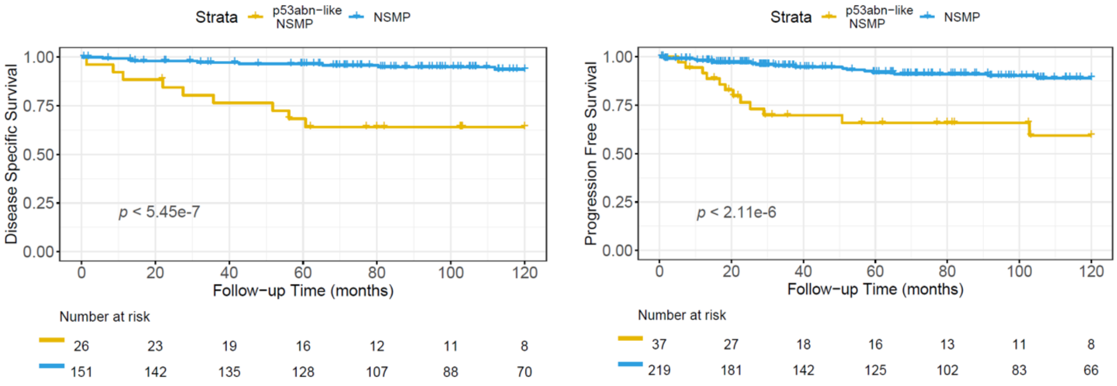

**B: Validation cohort**

**DSS** **PFS**

**Vanilla**

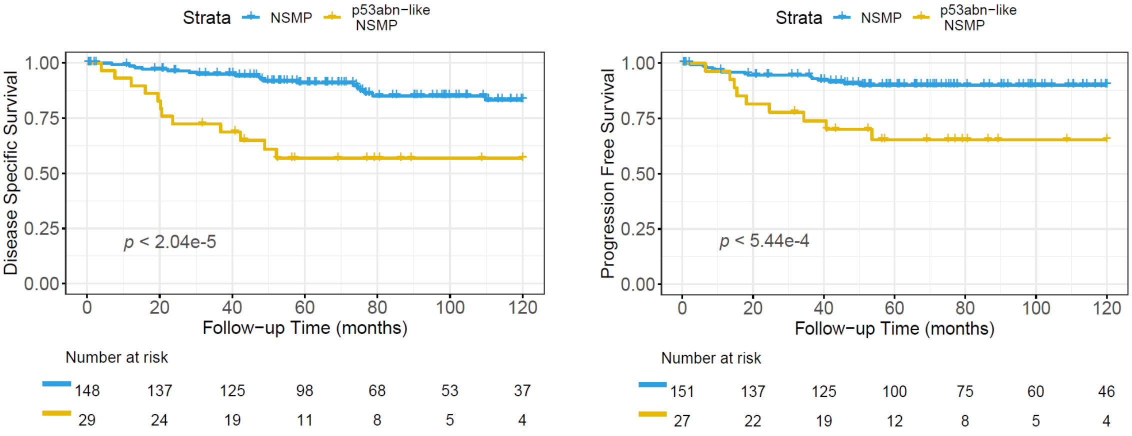

**IDaRS**

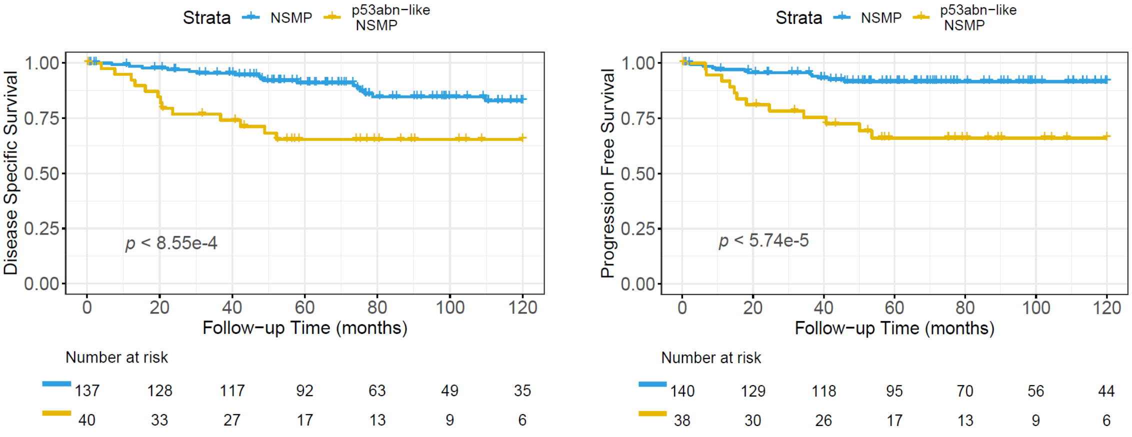

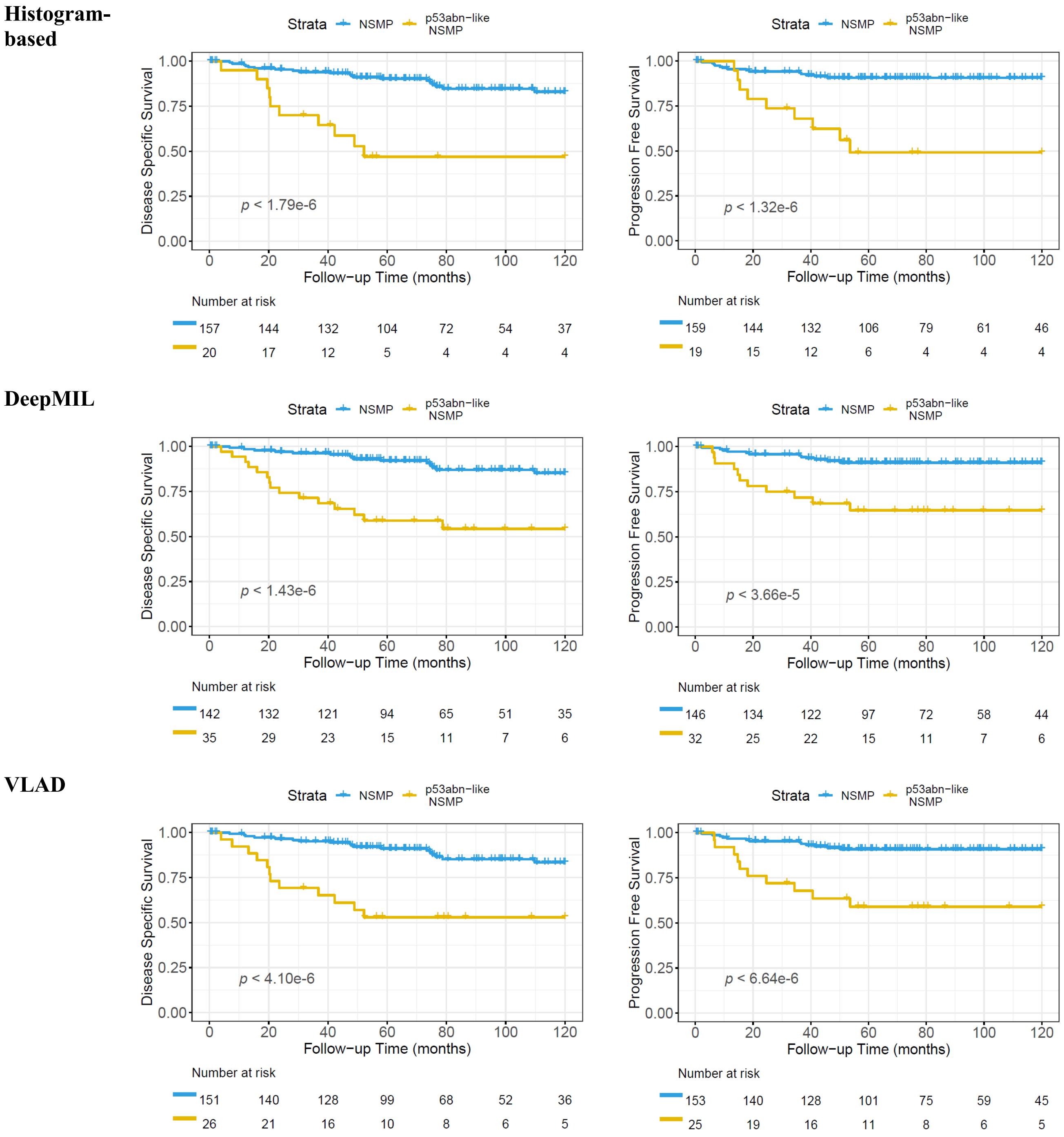

**Supplementary Figure 5:** Kaplan Meier curves along associated with the p53abn-like NSMP and NSMP groups for the discovery (A) and validation (B) cohorts using various deep learning frameworks. Note, DSS was not available for the TCGA part of the discovery cohort.

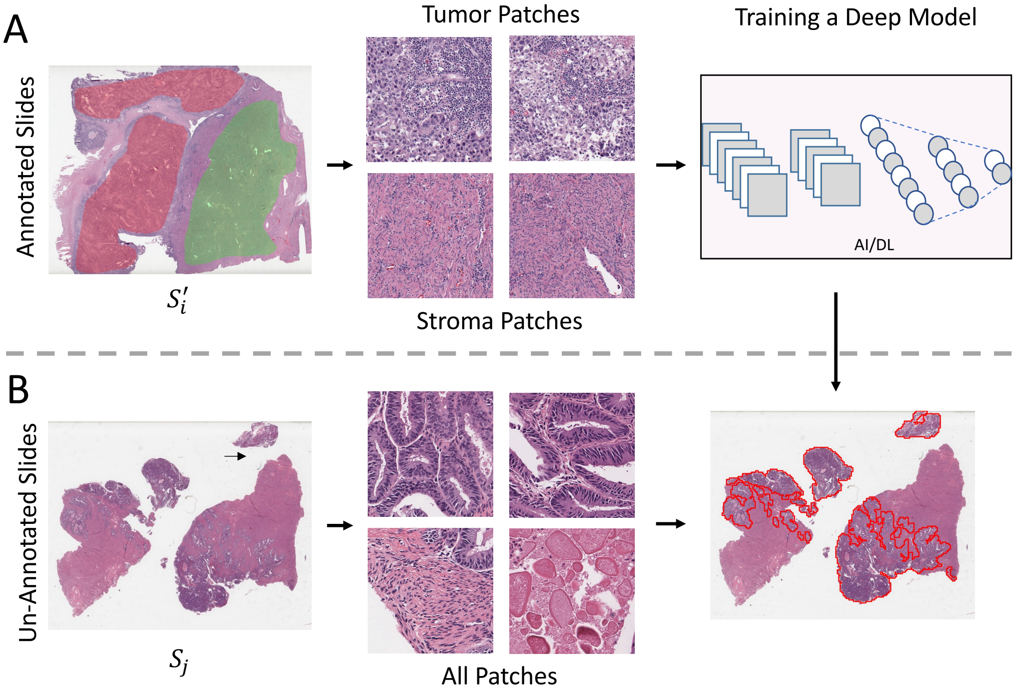

**Supplementary Fig. 6:** An overview of AI tumor-normal classifier and automatic annotation. (A) Extracting tumor and stroma patches from manually annotated slide and training a binary deep model. Red regions show annotated tumor regions, while green depicts stroma sections. (B) Extracting all patches from un-annotated slides and feeding them to the trained model to highlight only the tumor ones. The final tumor regions are shown by red contours.

**Supplementary Table 1:** Overview of cohorts.

| Dataset | Unit | Subtype | |
|---|---|---|---|
| | | NSMP | p53abn |
| TCGA (WSIs) | Patients | 90 | 56 |
| | Slides | 94 | 61 |
| | Manually Annotated Slides by Pathologists | 14 | 13 |
| | 512×512 Pixel Patches at 20x | 162,998 | 82,655 |
| German (WSIs) | Patients | 182 | 40 |
| | Slides | 355 | 76 |
| | Manually Annotated Slides by Pathologists | 0 | 0 |
| | 512×512 Pixel Patches at 20x | 576,553 | 125,376 |
| Vancouver (TMAs) | Patients | 195 | 95 |
| | Slides | 670 | 422 |
| | 512×512 Patches at 20x with 65% Overlap | 15,584 | 10,251 |

**Supplementary Table 2: Accuracy and other performance measures of the binary tumor-stroma classifier.**

| Non-tumor Accuracy | Tumor Accuracy | Kappa | F1 Score | AUC | Balanced Accuracy |
|---|---|---|---|---|---|
| 99·89% | 99·61% | 0·9952 | 0·9976 | 0·9994 | 99·75% |

**Supplementary Table 3:** Performance metrics of the deep learning model for p53abn vs. NSMP classifier. The results are based on mean ± std of 10 cross-validation splits.

| Source | p53abn Accuracy (%) | NSMP Accuracy (%) | Kappa | F1 score | AUC | Balanced Accuracy (%) |
|---|---|---|---|---|---|---|
| Discovery Set | 87·17 ± 10·86 | 91·59 ± 3·58 | 0·75 ± 0·09 | 0·87 ± 0·05 | 0·95 ± 0·03 | 89·38 ± 5·49 |
| Validation Set | 81·99 ± 4·56 | 77·61 ± 5·62 | 0·58 ± 0·03 | 0·79 ± 0·02 | 0·88 ± 0·01 | 79·80 ± 1·34 |

**Supplementary Table 4:** Detailed accuracy and other performance measures of the VarMIL network.

| Source | Split | Metrics | | | | | |
|---|---|---|---|---|---|---|---|
| | | p53abn Accuracy (%) | NSMP Accuracy (%) | Kappa | F1 score | AUC | Average Score (%) |
| Discovery Set | 1 | 70·97 | 90·48 | 0·6208 | 0·8104 | 0·9078 | 80·72 |
| | 2 | 64·52 | 89·41 | 0·5506 | 0·7752 | 0·9120 | 76·96 |
| | 3 | 96·00 | 93·33 | 0·8332 | 0·9164 | 0·9867 | 94·67 |
| | 4 | 92·59 | 91·67 | 0·7928 | 0·8961 | 0·9749 | 92·13 |
| | 5 | 96·30 | 85·71 | 0·7205 | 0·8586 | 0·9616 | 91·01 |
| | 6 | 96·30 | 89·41 | 0·7784 | 0·8885 | 0·9760 | 92·85 |
| | 7 | 96·15 | 87·76 | 0·7262 | 0·8617 | 0·9380 | 91·95 |
| | 8 | 88·46 | 94·90 | 0·8107 | 0·9053 | 0·9686 | 91·68 |
| | 9 | 80·77 | 97·98 | 0·8225 | 0·9112 | 0·9526 | 89·37 |
| | 10 | 89·66 | 95·29 | 0·8400 | 0·9200 | 0·9542 | 92·47 |
| Validation Set | 1 | 85·78 | 75·82 | 0·5897 | 0·7930 | 0·8836 | 80·80 |
| | 2 | 81·28 | 81·19 | 0·6121 | 0·8057 | 0·8822 | 81·24 |
| | 3 | 81·52 | 82·24 | 0·6263 | 0·8129 | 0·8972 | 81·88 |
| | 4 | 89·34 | 70·60 | 0·5619 | 0·7767 | 0·8869 | 79·97 |
| | 5 | 79·86 | 81·49 | 0·6032 | 0·8013 | 0·8892 | 80·68 |
| | 6 | 76·07 | 81·79 | 0·5733 | 0·7866 | 0·8766 | 78·93 |
| | 7 | 90·05 | 64·03 | 0·4974 | 0·7403 | 0·8679 | 77·04 |
| | 8 | 78·67 | 79·70 | 0·5722 | 0·7857 | 0·8755 | 79·19 |
| | 9 | 77·96 | 79·70 | 0·5660 | 0·7826 | 0·8731 | 78·83 |
| | 10 | 79·38 | 79·55 | 0·5767 | 0·7879 | 0·8742 | 79·47 |

**Supplementary Table 5:** Statistics of NSMP and *p53abn-like NSMP* patients based on p53abn vs. NMSP classifiers.

| Source | Number of patients | NSMP | *p53abn-like NSMP* |
|---|---|---|---|
| Discovery Set | 272 | 224 (82·35%) | 48 (17·65%) |
| Validation Set | 195 | 156 (80%) | 39 (20%) |

**Supplementary Table 6:** Performance benchmarking of Vanilla (A), IDaRS (B), Histogram (C), DeepMIL (D), and VLAD (E) models for the discovery and validation sets.

A: Vanilla

| Source | P53abn Accuracy (%) | NSMP Accuracy (%) | Kappa | F1 score | AUC | Balanced Accuracy (%) |
|---|---|---|---|---|---|---|
| Discovery Set | 74.80 ± 11.91 | 92.28 ± 5.51 | 0.68 ± 0.10 | 0.84 ± 0.05 | 0.93 ± 0.04 | 83.54 ± 5.69 |
| Validation Set | 79.46 ± 7.51 | 75.22 ± 11.43 | 0.53 ± 0.07 | 0.76 ± 0.04 | 0.84 ± 0.02 | 77.34 ± 2.85 |

B: IDaRS

| Source | p53abn Accuracy (%) | NSMP Accuracy (%) | Kappa | F1 score | AUC | Balanced Accuracy (%) |
|---|---|---|---|---|---|---|
| Discovery Set | 81.63 ± 9.78 | 88.61 ± 5.22 | 0.66 ± 0.07 | 0.83 ± 0.04 | 0.93 ± 0.03 | 85.12 ± 3.98 |
| Validation Set | 83.41 ± 5.21 | 71.19 ± 10.90 | & 0.52 ± 0.08 | 0.75 ± 0.05 | 0.84 ± 0.02 | 77.30 ± 3.34 |

C: Histogram

| Source | p53abn Accuracy (%) | NSMP Accuracy (%) | Kappa | F1 score | AUC | Balanced Accuracy (%) |
|---|---|---|---|---|---|---|
| Discovery Set | 72.38 ± 10.26 | 94.83 ± 2.64 | 0.70 ± 0.10 | 0.85 ± 0.05 | 0.88 ± 0.05 | 83.60 ± 5.34 |
| Validation Set | 75.88 ± 5.58 | 81.04 ± 4.95 | 0.56 ± 0.04 | 0.78 ± 0.02 | 0.80 ± 0.02 | 78.46 ± 1.77 |

D: DeepMIL

| Source | p53abn Accuracy (%) | NSMP Accuracy (%) | Kappa | F1 score | AUC | Balanced Accuracy (%) |
|---|---|---|---|---|---|---|
| Discovery Set | 85.72 ± 11.55 | 91.45 ± 4.31 | 0.74 ± 0.12 | 0.87 ± 0.06 | 0.95 ± 0.03 | 88.58 ± 6.60 |
| Validation Set | 82.84 ± 4.15 | 77.60 ± 5.65 | 0.59 ± 0.03 | 0.79 ± 0.02 | 0.88 ± 0.01 | 80.22 ± 1.32 |

E: VLAD

| Source | p53abn Accuracy (%) | NSMP Accuracy (%) | Kappa | F1 score | AUC | Balanced Accuracy (%) |
|---|---|---|---|---|---|---|
| Discovery Set | 79.11 ± 6.67 | 93.43 ± 2.45 | 0.73 ± 0.08 | 0.86 ± 0.04 | 0.92 ± 0.04 | 86.27 ± 3.87 |
| Validation Set | 77.48 ± 4.56 | 80.36 ± 3.50 | 0.57 ± 0.03 | 0.78 ± 0.01 | 0.84 ± 0.01 | 78.92 ± 1.53 |

**Supplementary Table 7:** Statistics of NSMP and p53abn-like NSMP patients based on p53abn vs. NMSP Vanilla (A), IDaRS (B), Histogram (C), DeepMIL (D), and VLAD (E) models for the discovery and validation sets.

A: Vanilla

| Source | Number of patients | NSMP | p53abn-like NSMP |
|---|---|---|---|
| Discovery Set | 272 | 213 (78.31%) | 59 (21.69%) |
| Validation Set | 195 | 163 (83.59%) | 32 (16.41%) |

B: IDaRS

| Source | Number of patients | NSMP | p53abn-like NSMP |
|---|---|---|---|
| Discovery Set | 272 | 200 (73.53%) | 72 (26.47%) |
| Validation Set | 195 | 152 (77.95%) | 43 (22.05%) |

C: Histogram

| Source | Number of patients | NSMP | p53abn-like NSMP |
|---|---|---|---|
| Discovery Set | 272 | 232 (85.29%) | 40 (14.71%) |
| Validation Set | 195 | 172 (88.21%) | 23 (11.79%) |

D: DeepMIL

| Source | Number of patients | NSMP | p53abn-like NSMP |
|---|---|---|---|
| Discovery Set | 272 | 228 (83.82%) | 44 (16.18%) |
| Validation Set | 195 | 157 (80.51%) | 38 (19.49%) |

E: VLAD

| Source | Number of patients | NSMP | p53abn-like NSMP |
|---|---|---|---|
| Discovery Set | 272 | 232 (85.29%) | 40 (14.71%) |
| Validation Set | 195 | 166 (85.13%) | 29 (14.87%) |

**Supplementary Table 8:** Clinicopathologic features of the *p53abn-like NSMP* group in the discovery set.

| Variable | Total | NSMP | *p53abn-like NSMP* | p53abn |
|---|---|---|---|---|
| **Total** | 363 | 221 (60·88%) | 47 (12·95%) | 95 (26·17%) |
| **Age at diagnosis** | | | | |
| <60 yrs | 121 (33·33%) | 90 (40·72%) | 20 (42·55%) | 11 (11·58%) |
| ≥60 yrs | 242 (66·67%) | 131 (59·28%) | 27 (57·45%) | 84 (88·42%) |
| **Histotype** | | | | |
| Endometrioid | 288 (79·34%) | 220 (99·55%) | 42 (89·36%) | 26 (27·37%) |
| Non-endometrioid | 75 (20·66%) | 1 (0·45%) | 5 (10·64%) | 69 (72·63%) |
| **Tumour grade** | | | | |

| | | | | |
|---|---|---|---|---|
| Low grade (G1–2) | 258 (71·07%) | 215 (97·29%) | 31 (65·96%) | 12 (12·63%) |
| High grade (G3) | 105 (28·93%) | 6 (2·71%) | 16 (34·04%) | 83 (87·37%) |
| **FIGO stage** | | | | |
| I-II | 291 (80·17%) | 203 (91·86%) | 36 (76·60%) | 52 (54·74%) |
| III-IV | 71 (19·56%) | 18 (8·14%) | 10 (21·28%) | 43 (45·26%) |
| Unknown | 1 (0·28%) | 0 | 1 (2·13%) | 0 |

**Supplementary Table 9:** Clinicopathologic features of the *p53abn-like NSMP* group in the validation set without excluding any patients.

| **Variable** | **Total** | **NSMP** | ***p53abn-like NSMP*** | **p53abn** |
|---|---|---|---|---|
| **Total** | 288 | 155 (53·82%) | 38 (13·19%) | 95 (32·99%) |
| **Age at diagnosis** | | | | |
| <60 yrs | 81 (28·13%) | 61 (39·36%) | 11 (28·95%) | 9 (9·47%) |
| ≥60 yrs | 205 (71·18%) | 92 (59·35%) | 27 (71·05%) | 86 (90·53%) |
| Unknown | 2 (0·69%) | 2 (1·29%) | 0 | 0 |
| **Histotype** | | | | |
| Endometrioid | 195 (67·71%) | 145 (93·55%) | 27 (71·05%) | 23 (24·21%) |
| Non-endometrioid | 91 (31·60%) | 8 (5·16%) | 11 (28·95%) | 72 (75·79%) |
| Unknown | 2 (0·69%) | 2 (1·29%) | 0 | 0 |
| **Tumour grade** | | | | |
| Low grade (G1–2) | 151 (52·43%) | 140 (90·32%) | 6 (15·79%) | 5 (5·26%) |
| High grade (G3) | 137 (47·57%) | 15 (9·68%) | 32 (84·21%) | 90 (94·74%) |
| **FIGO stage** | | | | |
| I-II | 216 (75·00%) | 138 (89·03%) | 28 (73·68%) | 50 (52·63%) |
| III-IV | 69 (23·96%) | 14 (9·03%) | 10 (26·32%) | 45 (47·37%) |
| Unknown | 3 (1·04%) | 3 (1·94%) | 0 | 0 |

**Supplementary Table 10:** Multi-variate Cox regression analysis showing the prognostic significance of *p53abn-like NSMP* group PFS for the entire cohort.

| **Variable** | **Hazard ratio** | ***p*** |
|---|---|---|
| Grade | 2·35 | 0·04 |
| Stage | 3·94 | 1.2e-5 |
| Histology (endometrioid vs. non-endometrioid) | 1.1 | 0.82 |
| *p53abn-like NSMP* | 2·53 | 0·01 |
